# Diagnostic Performance of Doppler Ultrasound for Acute Appendicitis: A Systematic Review and Diagnostic Test Accuracy Meta-Analysis

**DOI:** 10.1101/2025.05.01.25326795

**Authors:** Javier Arredondo Montero, María Rico-Jiménez

## Abstract

This systematic review and diagnostic test accuracy meta-analysis evaluated the performance of Doppler ultrasound (DUS) for diagnosing acute appendicitis (AA) and its ability to distinguish complicated (CAA) from non-complicated acute appendicitis (NCAA). The review was registered in PROSPERO (CRD42025641841), and a comprehensive search was conducted across PubMed, Web of Science, Scopus, and Ovid. Twenty-one studies were included. Study quality was assessed using the QUADAS-2 tool. Continuous quantitative parameters were synthesized using random-effects models (REML) while diagnostic performance data were analyzed through hierarchical models. For Spectral Doppler parameters, PSV showed a pooled sensitivity of 94% [89–97] and specificity of 87% [71–95], while RI showed a pooled sensitivity of 81% [68–89] and specificity of 88% [73–95]. Color Doppler (CD) achieved a pooled sensitivity of 82% [70–90], specificity of 97% [–99], and AUC of 0.97 [0.95–0.98], with borderline evidence of small-study effects (Deeks’ test p = 0.08). Evidence for discriminating CAA from NCAA was limited and heterogeneous. Although DUS—particularly CD and SD— shows promising diagnostic performance for AA, its accuracy is likely overestimated due to moderate heterogeneity, retrospective designs, small-study effects (CD), and spectrum bias. Still, the consistently high estimates across modalities, combined with its radiation-free nature and wide availability, underscore its value as a complementary imaging tool. Until confirmed by large, prospective, multicenter studies with standardized protocols, DUS should be viewed as a promising option—especially in pediatric and radiation-sensitive populations—when the appendix is adequately visualized.

**Funding:** None.

**Registration:** PROSPERO (CRD42025641841).

## Introduction

Acute appendicitis (AA) remains the leading cause of emergency abdominal surgery worldwide [1]. Although its overall mortality rate is low, diagnostic delays significantly increase the risk of complications and morbidity [2,3].

AA diagnosis is primarily based on physical examination, a focused clinical history, and basic laboratory tests, including complete blood count and acute-phase reactants. The current recommended approach emphasizes risk stratification to guide clinical decision-making, employing multivariable scoring systems such as the AIR score, the RIPASA score, the PAS score, and the BIDIAP score—the latter two specifically developed for pediatric populations—to identify patients who require additional imaging and/or hospital admission, and to reduce the incidence of negative surgical explorations [4–6]. Although these scoring systems have proven highly effective for the initial triage of patients with suspected AA, imaging techniques— primarily ultrasound (US) and computed tomography (CT)—remain essential for confirming or ruling out the diagnosis and for differentiating between complicated (CAA) and uncomplicated (NCAA) appendicitis [7,8].

Computed tomography (CT) is a widely recognized diagnostic tool for AA, particularly in cases with a high clinical suspicion and inconclusive US findings, having shown superior diagnostic performance compared to US in recent meta-analyses [7]. However, using CT involves considerable expenditure of human and economic resources. In addition, despite ongoing advances in low-dose imaging protocols, CT remains a significant source of ionizing radiation, which limits its unrestricted use, particularly in vulnerable populations such as children and pregnant women [1,4,7]. Despite the available evidence, recent studies continue demonstrating an overuse of CT imaging in pediatric populations [9].

Other imaging modalities, such as magnetic resonance imaging (MRI), have also demonstrated excellent diagnostic performance in AA [10,11]. However, their clinical implementation remains challenging and costly in current practice. For instance, pediatric patients often require sedation to undergo MRI examinations, adding complexity to its routine use.

US has demonstrated excellent diagnostic performance in the evaluation of appendicitis, both when performed by specialized radiologists [7] and when conducted by clinicians using point-of-care ultrasound (POCUS) [12,13]. Nevertheless, US remains a highly operator-dependent modality, and considerable rates of non-visualization of the cecal appendix are reported in recent literature [14]. Non-visualization may be attributed to several factors, including patient obesity, the anatomical location of the appendix, poor acoustic windows due to interposed bowel loops, and the operator’s experience level. The adoption of standardized protocols, such as the graded compression technique described by Puylaert in 1986 [15], the three-step positioning algorithm [16], and structured coaching strategies [17], has significantly improved appendiceal visualization rates, but they remain far from perfect.

Quillin et al. first reported using Doppler US (DUS) as an additional diagnostic tool in evaluating AA in 1992 [18]. Based on the pathophysiological premise that inflammation of the cecal appendix leads to increased blood flow that can be detected and quantified using Doppler techniques, numerous studies have evaluated the potential diagnostic performance of DUS— including color Doppler (CD), power Doppler (PD), contrast-enhanced power Doppler (CEPD), and more recently, spectral Doppler (SD)—in AA, as well as its ability to discriminate between CAA and NCAA [19–38]. However, the available evidence remains fragmented: most studies are small, single-center, and heterogeneous in design, and previous reviews have not provided a quantitative synthesis of DUS performance across modalities. In particular, little is known about whether DUS can aid in discriminating CAA from NCAA, a clinically relevant distinction that influences urgency and management strategy. This systematic review and DTA meta-analysis addresses these gaps by providing a modality-specific quantitative synthesis and exploring discrimination between CAA and NCAA.

## Methods

### Literature search and selection

We followed the Preferred Reporting Items for Systematic Reviews and Meta-Analyses in Diagnostic Test Accuracy Studies (PRISMA-DTA) guidance [39]. Supplementary File 1 shows the PRISMA-DTA Checklist. We prospectively registered the present review in the International Prospective Register of Systematic Reviews (PROSPERO ID CRD42025641841).

Eligible studies were identified by searching the primary existing medical bibliography databases (PubMed, Web of Science, Scopus, and Ovid). Supplementary File 2 shows the detailed search strategy for each bibliographic database. The search was last executed on 22.04.2025.

JAM and MRJ selected articles using the COVIDENCE ® tool. The search results were imported into the platform, and both authors screened the articles separately. Disagreements were resolved by consensus. We included prospective or retrospective observational clinical studies evaluating the diagnostic performance of any Doppler ultrasound modality (e.g., CD, PD, or SD) for acute appendicitis in adult or pediatric populations, with histopathology of the resected appendix as the reference standard. We excluded case reports, reviews, duplicate or overlapping datasets, retracted publications, studies in languages other than English or Spanish, and those not involving surgical confirmation or populations outside the scope of interest (e.g., immunocompromised patients, abdominal malignancy, hematological disorders). Supplementary File 3 shows the inclusion and exclusion criteria.

### Quality assessment

The QUADAS-2 (Quality Assessment of Diagnostic Accuracy Studies 2) tool was used to evaluate each selected article’s methodological quality and risk of bias [40]. Each article evaluated patient selection, index test, reference standard, flow, and timing. Applicability concerns regarding patient selection, index tests, and reference standards were also assessed. Two authors independently performed the assessments, and disagreements were resolved by consensus. As no conflicts remained unresolved, a third reviewer was not required.

### Data extraction and synthesis

The target condition was defined as AA confirmed either by histopathological examination or intraoperative findings. The index test was DUS (all modes). The reference standard was the histopathological examination of the resected cecal appendix. Two independent reviewers (JAM, MRJ) extracted the relevant data from the selected articles following a standardized procedure. Extracted data included author, country where the study was conducted, year of publication, study design, study population (sample size, age range, and sex distribution), AA group and control group (CG) definitions, reference standard used in AA group, mean or median and standard deviation or range or interquartile range for peak systolic velocity (PSV) and resistive index (RI) determinations, statistical p-value for the between-group comparison, PSV and RI cut-off value (if established), and its associated sensitivity and specificity. There were no disagreements between the reviewers after collating the extracted data. The metrics used in each study were reviewed, and it was determined that a standardization of units was not required. Means (ranges) were converted to means (standard deviations) following a standardized procedure [41,42] in two cases [33,38]. True positives (TP), false positives (FP), true negatives (TN), and false negatives (FN) were obtained either directly from the included studies or estimated, when not explicitly reported, based on available sensitivity, specificity, and the number of patients with and without the target condition, using standardized statistical formulae [43]. Reported sensitivities, specificities, sample sizes per group, and predictive values were used to cross-validate the calculations.

The diagnostic odds ratio (DOR) was calculated for each study as (TP × TN) / (FP × FN). A continuity correction of 0.5 was applied to all zero-valued contingency table cells to avoid division by zero. Subsequently, DOR was log-transformed to stabilize variances and allow for linear approximation. The log-transformed DOR’s standard error (SE) was calculated using standard formulae based on the corrected contingency table counts. The 95% confidence intervals (CIs) for the log(DOR) were obtained by applying the normal approximation method (log(DOR) ± 1.96 × SE). They were then exponentiated to derive the 95% CIs on the original DOR scale.

### Random-Effects Meta-analysis

Two random-effects meta-analyses (MAs) for DUS quantitative parameters were performed using the restricted maximum likelihood (REML) method: (1) a meta-analysis of PSV values (measured in cm/s) comparing AA and CG patients, and (2) a meta-analysis of RI values comparing AA and CG patients. Confidence intervals (CIs) were adjusted using the truncated Hartung–Knapp–Sidik–Jonkman (t-HKSJ) approach whenever the number of studies (*k*) was greater than two and the between-study variance (τ²) was non-zero.

These analyses, which did not formally evaluate diagnostic performance, were conducted in an exploratory manner, aiming to obtain numerical reference values with potential clinical applicability. All studies with available data were included. Due to the limited number of studies reporting PSV and RI, no additional sensitivity analyses were conducted. Results were expressed as mean differences with corresponding 95% confidence intervals (CIs) and 95% prediction intervals (PIs) and were depicted using forest plots. Between-study heterogeneity was assessed using Cochran’s Q test, the between-study variance (τ²), and the inconsistency index (I²). Two leave-one-out sensitivity analyses were conducted (one for each meta-analysis).

### Diagnostic Test Accuracy Meta-analysis

Four main hierarchical (DTA) meta-analytical models were conducted: (1) overall diagnostic performance of DUS (AA versus CG), (2) diagnostic performance of SD (PSV modality), (3) diagnostic performance of SD (RI modality) (AA versus CG), and 4) diagnostic performance of CD (AA versus CG).

In the overall DUS model (1), different Doppler modalities from the same study populations were combined, meaning that some studies contributed more than one observation. This lack of statistical independence can reduce estimated variance and artificially inflate precision. Therefore, the overall model is presented strictly as exploratory and descriptive, while confirmatory inferences rely on the modality-specific models (CD and SD). Pooled sensitivity, specificity, and area under the curve (AUC) estimates were reported for each model. Results were presented as forest plots of sensitivity and specificity and hierarchical summary receiver operating characteristic (HSROC) curves. Pre-specified meta-regression analyses were conducted on the primary model to explore the influence of study design (prospective vs. retrospective) and population characteristics (pediatric vs. mixed/adult) on diagnostic performance. The *metadta*, *midas*, and *metandi* modules in STATA were used to conduct the DTA meta-analyses [44–46]. The *mada* module in R and the *midas* module in STATA were used for the meta-regression DTA analyses [47].

### Fagan nomogram

A Fagan nomogram was constructed to assess the clinical utility of DUS by estimating post-test probabilities based on pooled likelihood ratios in CD and SD DTA models. A pretest probability of 20% was selected to reflect a typical clinical scenario of intermediate suspicion for acute appendicitis in Emergency settings. Pooled positive and negative likelihood ratios (LR and LR ) derived from the hierarchical meta-analysis were applied to calculate post-test probabilities following positive and negative test results.

### Publication Bias and Small-Study Effects Assessment

For the random-effects meta-analytical models, Egger’s and Begg’s tests, funnel plots, and the trim-and-fill method were initially considered to assess the risk of publication bias [48]. However, since neither model included a sufficient number of studies (n < 10), these analyses were not performed. For the DTA meta-analytical models, Deeks’ asymmetry test was performed when more than 10 studies were included in the analysis to evaluate the presence of publication bias [49]. A p-value <0.10 was considered suggestive of publication bias, in line with established guidelines.

Statistical analyses were conducted using Stata version 19.0 (StataCorp LLC, College Station, TX, USA) with the *metandi*, *midas,* and *metadta* modules, and R version 4.3.2 (R Foundation for Statistical Computing, Vienna, Austria) with the *mada* module (version 0.5.12).

## Results

The search returned 405 articles (Scopus, n = 92; PubMed, n = 91; Web of Science, n = 192; Ovid MEDLINE, n = 30). One hundred seventeen duplicates were removed. Among the remaining 288 articles, we excluded 267 (inclusion and exclusion criteria, n=267; reports not retrieved, n=0). This review finally included 21 studies [18–38]. A total sample size, as well as specific counts per group (AA/CG and gender), could not be reliably provided. This is due to the identification of multiple discrepancies across studies, which often lacked explicit reporting of these figures. The flowchart of the search and selection process is shown in Figure 1.

**Figure 1.**
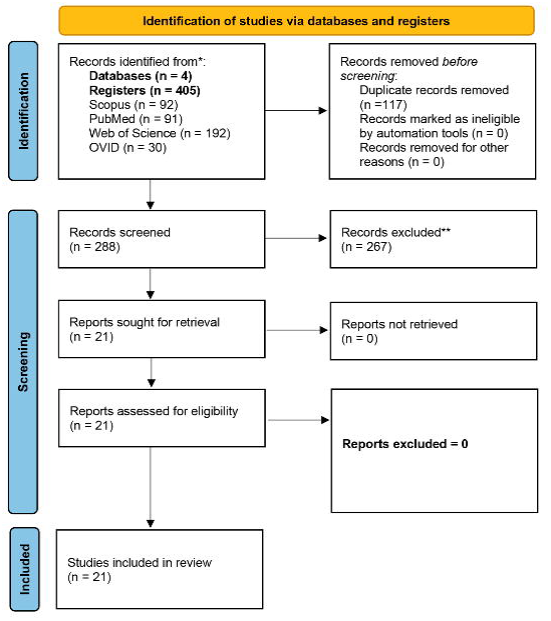
Flowchart of the search and selection process.

The risk of bias concerning the selection of patients was considered low in three of the studies [18–20], unclear in 17 of them [21–23,25–29,31–38], and high in two of them [24,30]. The risk of bias concerning the index test was considered low in 11 studies [18,21–28,30,32], unclear in six [19,20,22,29,31,34], and high in five [33,35–38]. The risk of bias concerning the reference standard was considered low in four studies [20,31,32,34], unclear in 17 [19,21–30,33,35–38], and high in one of them [18]. The risk of bias concerning flow and timing was considered low in 15 studies [18,19,21–28,32,35–38], unclear in six studies [20,22,29,31,33,34], and high in one of them [30]. Regarding patient selection applicability concerns, the risk was considered low in three of the studies [18–20], unclear in 17 of them [21–23,25–29,31–38], and high in two of them [24,30]. Regarding the index test applicability concern, the risk was considered low in 18 studies [18,20–30,32,35–38] and unclear in four studies [19,31,33,34]. Concerning reference standard applicability concerns, the risk was considered low in 20 studies [19–38] and high in one [18]. In the case of Lim et al. [22], prospective and retrospective cohorts were analyzed separately. For this reason, the reference may be considered as both low risk and unclear risk in certain categories (such as index test or flow and timing). The QUADAS-2 results are depicted in Figure 2.

**Figure 2.**
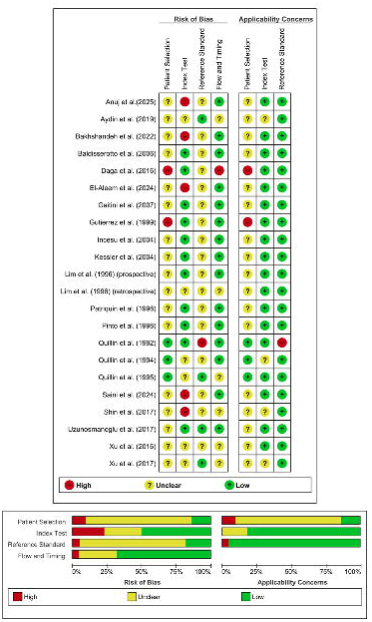
Graphical representation of the quality assessment of the diagnostic accuracy studies included in the review (QUADAS-2 tool).

### Doppler Ultrasound in Acute Appendicitis

#### Sociodemographic Characteristics

Table 1 summarizes the data extracted from the twenty-one studies that evaluated DUS. All studies were conducted between 1992 and 2025 [18–38]. Seven were from the United States [18–20,24,29,31,33], three were from India [30,37,38], three were from Turkey [26,32,34], one was from Canada [21 ], one was from South Korea [22], one was from Italy [23], one was from France [25], one was from Brazil [27], one was from Israel [28], one was from Iran [35], one was from Egypt [36]. Thirteen studies were prospective [18–21,23–27,32,36–38], and five were retrospective [28,29,31,33,34]. One study reported two cohorts, one prospective and one retrospective [22]. One study did not explicitly report its design; after reviewing it, we classified it as retrospective [30]. One study was reported as cross-sectional, and after reviewing its design, we classified it as retrospective [35]. Four studies involved exclusively pediatric populations [18–20,27].

Fifteen studies included patients with clinical suspicion of AA as their study population [18–20, 23, 25–30, 33, 35–38]. One study included a selective group of patients presenting with atypical manifestations of AA [24]. In two cases, populations with histopathologically confirmed AA and various types of CG were included separately [21, 22]. Three studies included only patients who underwent surgical intervention for suspected AA [31, 32, 34].

Twenty studies consistently defined ’case’ as the histopathological confirmation of AA in the surgical specimen [19–38]. AA was based on surgical findings in one study, and a histopathological study was not explicitly reported [18]. Twelve studies stratified the AA group into NCAA and CAA [18–21,23–28,31,32]

This was not the case for the definition of ’control’, which constituted either patients seen at the Emergency Department in which the diagnosis of AA was finally excluded (also known as non-surgical abdominal pain or NSAP) [18,22–30,33,34], patients with AA suspicion which finally had other surgical pathology [18,24,28], negative appendectomies (NA) [19,23,24,26–30,32] or specifically lymphoid hyperplasia as a form of NA [34], healthy control with ultrasound performed for other reasons (i.e, urological pathology) [21], patients with irritable bowel syndrome suspicion who underwent a barium enema [22].

In 12 studies, the authors restricted their analyses to the subgroup of patients in whom the cecal appendix was identified using grayscale ultrasound (US) [21–23,25,27–30,33,35,36,38]. Additionally, in some of these cases, the inclusion criteria were even more restrictive. For instance, in the study by Daga et al., only the 85 patients with appendiceal identification on US and sonographic criteria for acute appendicitis were included [30], while in the study by Anuj et al., only patients with an appendix visible on grayscale US and spectral Doppler waveforms on appendiceal US were considered [38]. In three of these cases, only US examinations with borderline features were included [22, 29, 35].

Table 1 shows the main characteristics of the studies included in this review, including the Doppler modalities assessed and the technical parameters of the sonographic examinations.

### Overall Doppler Ultrasound Diagnostic Performance in Acute Appendicitis

#### Exploratory Diagnostic Test Accuracy Meta-Analysis for Overall Doppler Modalities (AA versus CG, non-independent)

In the case of Baldisserotto et al., the most favorable TP, FP, TN, and FN data reported by the authors were used, based on the classification of appendiceal Doppler flow adopted in each study (see Youden’s J index value in Table 1). Daga et al. were excluded from the DTA meta-analyses because the diagnostic performance data reported included only patients with AA, and the manuscript contained numerical inconsistencies (see Table 1).

The DTA meta-analysis for overall Doppler modalities, including CD, PD, CEPD, and SD (AA versus CG), included 26 observations from 21 studies and yielded a pooled sensitivity and specificity [95% CI] of 86% [79–91] and 94% [90–96], respectively (Supplementary File 4). The pooled area under the ROC curve (AUC) [95% CI] was 0.96 [0.94-0.97]. Between-study heterogeneity (τ²) was 1.06 for sensitivity and 1.48 for specificity, with a negative correlation (ρ = –0.36) between them.

This pooled DUS model combined multiple modalities from the same cohorts; observations are therefore non-independent, which may reduce variance and inflate precision. Accordingly, this analysis should be regarded strictly as exploratory, intended to provide an approximate overview of DUS performance across modalities rather than definitive evidence.

We performed prespecified univariable meta-regressions to explore whether study design (prospective versus retrospective) or population type (pediatric versus mixed/adult) influenced the diagnostic performance of DUS. Each covariate was analyzed separately. No statistically significant association was found in the joint models. However, prospective studies showed significantly lower specificity (93% versus 95%; p = 0.01), and pediatric studies showed a non-significant trend toward higher sensitivity (93% versus 85%; p = 0.13). Deeks’ funnel plot asymmetry test did not indicate significant small-study effects (slope = 7.31, p = 0.34).

Supplementary File 5 includes the raw TP, FP, FN, and TN dataset used for all DTA analyses. It also includes the DOR (95% CI) calculated for each study.

### Spectral Doppler

#### Spectral Doppler Measurement Units

Nine studies reported the use of SD for diagnosing acute appendicitis (AA) [21,22,29,32,33,35,36–38]; among them, seven provided numerical values and/or specific diagnostic performance data [21,32,33,35–38]. The authors who reported SD numerical values and/or diagnostic performance data assessed it using three continuous quantitative parameters: PSV, RI, and pulsatility index (PI). PSV was consistently reported across all studies in centimeters per second (cm/s), whereas RI is a dimensionless parameter.

In two instances, where studies reported means and ranges instead of standard deviations, the missing standard deviations were estimated using the method described by Wan et al. [41] to enable meta-analytic pooling. Although the Wan et al. method was initially developed to estimate means and standard deviations from medians and ranges (or interquartile ranges), the studies in question reported means (not medians). To minimize potential inaccuracies, results were compared with estimates obtained using the method proposed by Hozo et al. [42]. In addition, an exploratory assessment of skewness following the approach of Shi et al. did not suggest that the underlying distributions were formally skewed. Therefore, conversions based on quantiles, such as those proposed by McGrath et al., were not applied. Nevertheless, this approach constitutes a methodological limitation, as combining means and ranges alone does not reliably allow accurate estimation of standard deviation and may not accurately reflect the underlying data distribution.

#### Diagnostic Performance of Peak Systolic Velocity and Resistive Index (AA Versus CG)

Seven studies reported quantitative values of the RI [21,26,32,33,36,37,38]. Five of them [33,35–38] reported a cut-off value for RI, ranging from 0.495 [35] to 0.65 [33,38]. Sensitivities and specificities for RI ranged from 63.9% [33] to 90.5% [35] and from 58.3% [37] to 96.5% [33], respectively.

Four studies reported quantitative values in cm/s for PSV [33,36,37,38]. They all reported a PSV cut-off, ranging from 8.6 cm/s [36] to 11.8 cm/s [37]. Sensitivities and specificities for PSV ranged from 85.3% [38] to 98.3% [36] and from 54.2% [37] to 94.7% [33,35], respectively. One study reported PI values as means [32]. In five studies, true positive, false positive, true negative, and false negative values could be calculated for both PSV and RI [33,35,36,37,38].

Four studies provided a p-value for the comparison of PSV and RI values between the AA and CG, three of which were statistically significant (p<0.001) [33,36,38]. In the study by Saini et al. [37], the p-value for the comparison of PSV between groups was statistically significant (p<0.009). In contrast, the p-value for the comparison of RI only reached marginal significance (p=0.056). The reported sensitivity and specificity for each study are shown in Table 1.

#### Random-Effects Meta-Analysis for Spectral Doppler (AA Versus CG)

The random-effects meta-analysis of PSV (AA versus CG) included four studies (139 AA patients and 139 CG) and showed a significant mean difference [95% CI] of 7.43 [5.37–9.48] cm/s (p = 0.01). Cochran’s Q test yielded a χ² value of 7.62, I² indicated moderate heterogeneity (59.6%), and the between-study variance (τ²) was 2.53. After applying the t-HKSJ adjustment, the 95% CI was [4.08–10.77] cm/s. The 95% PI was [-0.77,15.63] cm/s. The forest plot of this meta-analysis is shown in Figure 3 (upper panel).

**Figure 3.**
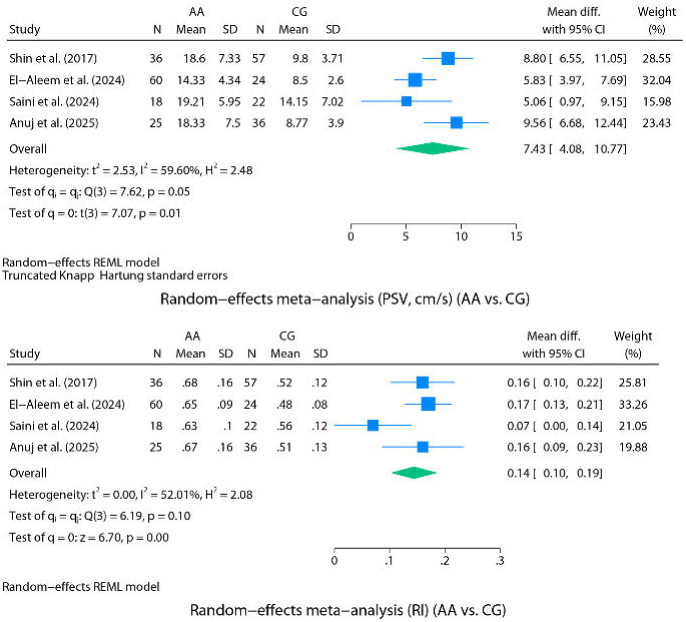
<u>Above</u>: Forest plot of the REML random-effects meta-analysis performed for PSV (cm/s) (AA versus CG). <u>Bottom</u>: Forest plot of REML random-effects meta-analysis performed for RI (AA versus CG).

A leave-one-out analysis was performed, carrying out iterations on the present model, excluding one of the studies included in each iteration (forest plot not shown). The leave-one-out analysis revealed that the article that most negatively conditioned the model was El-Aleem et al. [36]. Its exclusion from the model resulted in a mean difference [95% CI] of 8.34 [6.42-10.27] cm/s (p<0.001). Given that fewer than 10 studies were included in the model, no formal assessment of publication bias was conducted, and no trim-and-fill imputation methods were applied, in line with current methodological recommendations.

Concerning the random-effects meta-analysis of RI, Patriquin et al. [21] reported CG RI values as a range without a measure of central tendency; thus, the study could not be included in the meta-analytical models. Incesu et al. [26] did not provide a dispersion measure for RI, and Uzunosmanoğlu et al. (2017) [32] likewise reported RI values without dispersion data; consequently, both studies were also excluded from the meta-analyses. The random-effects meta-analysis of RI (AA versus CG) included four articles (139 AA and 139 controls) and yielded a significant mean difference [95% CI] of 0.14 [0.10-0.19] (p < 0.01). Cochran’s Q test yielded a χ² value of 6.19, I² indicated moderate heterogeneity (52%), and the between-study variance was τ² = 0.00. This discrepancy between I² and τ² can occur in meta-analyses with a small number of studies because the model has insufficient statistical power to distinguish true between-study differences from random sampling error reliably. In this case, no t-HKSJ adjustment was applied. The 95% PI was [-0.02, 0.31]. The forest plot of this meta-analysis is shown in Figure 3 (lower panel).

A leave-one-out analysis was performed, carrying out iterations on the present model, excluding one of the studies included in each iteration (forest plot not shown). The leave-one-out analysis revealed that the article that most negatively conditioned the model was Saini et al. [37]. Its exclusion from the model resulted in a mean difference [95% CI] of 0.17 [0.13-0.20](p<0.001). Given that fewer than 10 studies were included in the model, no formal assessment of publication bias was conducted, and no trim-and-fill imputation methods were applied, in line with current methodological recommendations.

#### Diagnostic Test Accuracy Meta-Analysis for Spectral Doppler (AA versus CG)

The model for PSV (AA versus CG) included five studies and yielded a pooled sensitivity and specificity [95% CI] of 94% [89–97] and 87% [71–95], respectively (Figure 4). Between-study heterogeneity (τ²) was 0.29 for sensitivity and 1.05 for specificity, with a perfect negative correlation between them (ρ = –1.00). Such a perfect correlation is unusual and most likely reflects model instability due to the small number of studies rather than a reliable estimate. A Fagan nomogram was used with a pretest probability of 20%. The post-test probability increased to 63.6% after a positive result (LR = 7) and decreased to 5.2% after a negative result (LR = 0.22).

**Figure 4.**
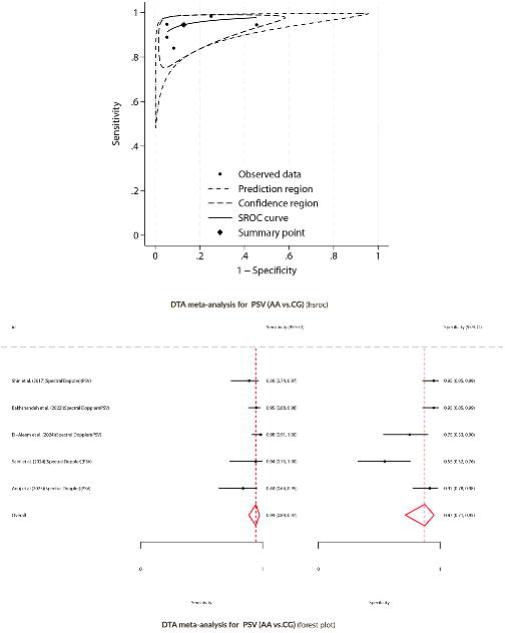
<u>Above</u>: DTA meta-analysis for PSV (AA versus CG). HSROC curve. <u>Bottom</u>: Forest plot of the DTA meta-analysis performed for PSV (AA versus CG).

The model for RI (AA versus CG) included five studies and yielded a pooled sensitivity and specificity [95% CI] of 81% [68–89] and 88% [73–95] (Figure 5). Between-study heterogeneity (τ²) was 0.41 for sensitivity and 0.91 for specificity, with a strong negative correlation between them (ρ = –0.91). A Fagan nomogram was used to estimate post-test probabilities based on a pretest probability of 20%. The post-test probability increased to 62% following a positive result (LR = 7) and decreased to 5% after a negative result (LR = 0.22).

**Figure 5.**
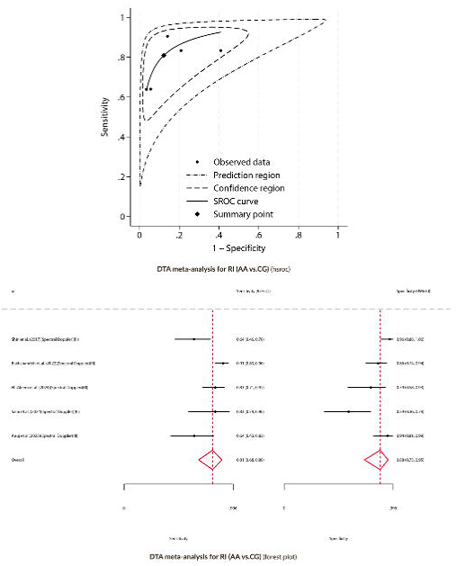
<u>Above</u>: DTA meta-analysis for RI (AA versus CG). HSROC curve. <u>Bottom</u>: Forest plot of the DTA meta-analysis performed for RI (AA versus CG).

### Color Doppler

Fifteen authors evaluated CD as a diagnostic tool in acute appendicitis (AA) [18–25,27–32,34]. Of these authors, two exclusively assessed the ability of CD to discriminate between NCAA and CAA [20,31], while the rest evaluated the ability of CD to diagnose AA in comparison to the CG [18,19,21–25,27–30,32,34]. A considerable heterogeneity was identified in the reported definitions of positivity (pathological findings) for CD imaging in acute appendicitis (AA). While some authors considered any detection of CD flow in the cecal appendix as positive, others only considered positivity when hyperemia or increased appendiceal flow was observed. Some authors, such as Patriquin et al.[21], used a multicategory scale based on the number of CD signals detected in the appendiceal wall (0 = none, 1–2 = few, 3–4 = moderate, >4 = abundant). This scale was later replicated by other authors, such as Gaitini et al. [28]. Some studies reported different diagnostic performance estimates depending on the cut-off point selected for the proposed scale; for example, Xu et al.[29] reported varying results depending on whether elevated flow or "type 2 flow" was considered diagnostic of AA, or whether the absence of flow was deemed sufficient to exclude AA. Other authors, such as Daga et al.[30], also reported different diagnostic outcomes depending on whether any detected appendiceal Doppler flow was considered diagnostic, or only cases showing hyperemia.

#### Diagnostic Test Accuracy Meta-Analysis for Color Doppler (AA versus CG)

The DTA meta-analysis for CD (AA versus CG) included 13 independent observations from 12 studies and yielded a pooled sensitivity and specificity [95% CI] of 82 [70–90] % and 97 [92–99] % (Figure 6). Between-study heterogeneity was moderate to substantial. τ² was 1.04 for sensitivity and 1.39 for specificity. The correlation between sensitivity and specificity was weakly negative (ρ = –0.11). The pooled AUC [95% CI] was 0.97 [0.95-0.98]. Visual inspection of Deeks’ funnel plot and the results of the asymmetry test (p = 0.08) did suggest the presence of potential publication bias among the included studies (figure not shown). The potential presence of publication bias suggests that the pooled sensitivity and specificity estimates for CD should be interpreted with caution, as they likely represent the most optimistic scenario of the available evidence. A Fagan nomogram was used to estimate post-test probabilities based on a pretest probability of 20%. The post-test probability increased to 86% following a positive DUS result (LR = 24) and decreased to 4% after a negative result (LR = 0.19). These findings support DUS’s strong rule-in and moderate rule-out value in clinical scenarios with intermediate pretest suspicion.

**Figure 6.**
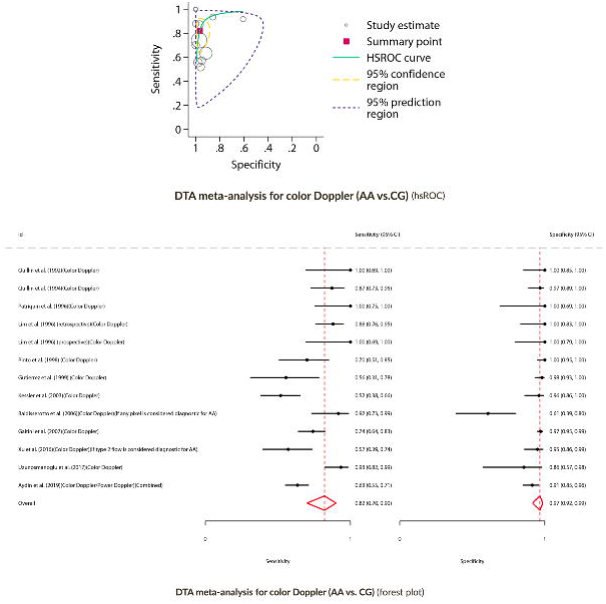
<u>Above</u>: DTA meta-analysis for color Doppler (AA versus CG). HSROC curve. <u>Bottom:</u> Forest plot of the DTA meta-analysis performed for color Doppler (AA versus CG).

Lastly, a *post-hoc* exploratory DTA sensitivity analysis was performed to address a discrepancy identified during data extraction for the Aydin et al. study. This analysis, which excluded the said study to assess its impact on the pooled estimates, showed a pooled sensitivity of 84% [95% CI: 71–91] and a specificity of 97% [95% CI: 92–99]. These values do not substantially differ from those obtained in the main model, which included 13 observations.

### Power Doppler

Three authors reported evaluating PD to diagnose acute appendicitis (AA) [23,26,34]. Pinto et al. reported a higher diagnostic performance of PD over CD [23]. Incesu et al. PD with CEPD, demonstrating the latter’s superiority over standalone PD [26]. Aydin et al. reported diagnostic performance data that combined results from both CD and PD modalities without distinction [34].

Only three studies’ contingency table data (TP, FP, TN, FN) were available for the power Doppler modality. Therefore, a DTA meta-analytical model could not be performed, as at least four studies are required to fit such models reliably.

### Doppler Ultrasound (Complicated Appendicitis versus Non-Complicated Appendicitis)

Four studies provided Doppler data and/or comparisons for CAA and NCAA groups [20,21,31,32].

#### Diagnostic Performance of Doppler Ultrasound (NCAA versus CAA)

Four studies reported the sensitivity and specificity of CD for discriminating NCAA and CAA: Quillin et al. [20] (77.8% and 60%), Patriquin et al. [21] (100% for both), Uzunosmanoğlu et al. [32] (93% and 85%), and Xu et al. [31] (25% and 72.4%). Two studies also provided SD measurements for the CAA and NCAA groups, using RI values [21] or PI values [32].

## Discussion

The present systematic review and meta-analysis evaluated the role of all Doppler US modalities in diagnosing AA, consistently demonstrating excellent diagnostic yield.

Concerning the biological plausibility and the pathophysiological rationale for using DUS to diagnose AA, inflammation of the cecal appendix is associated with a localized increase in vascular perfusion secondary to the release of inflammatory mediators. These changes can be potentially detected through Doppler imaging techniques. However, it is essential to note that this phenomenon is not specific to AA and may occur in any infectious or inflammatory process. Consequently, conditions such as colitis or ileitis may also present with increased Doppler signal on ultrasound evaluation. However, it should be considered that based on this same pathophysiological premise, the occurrence of appendiceal tissue ischemia in the context of gangrenous acute appendicitis (GAA) or CAA may be associated with a reduction or absence of Doppler flow within the appendix. This phenomenon has been previously reported by authors such as Quillin et al. [20], who observed that appendiceal hyperemia was more frequent in non-perforated AA compared to perforated cases, and Patriquin et al. [21], who described the absence of a Doppler signal at the appendiceal tip in a high proportion of CAA cases.

Regarding the different Doppler modalities, CD, PD, and SD have been primarily evaluated. CD was the first modality used for diagnosing AA and remains the most extensively characterized in the medical literature, demonstrating strong diagnostic yield. PD has also shown excellent, and in some cases superior, performance; however, the limited number of published studies and the inability to conduct meta-analytical models to assess its diagnostic accuracy quantitatively prevent definitive conclusions from being drawn. CEPD, although promising, was evaluated in only one article. On the other hand, recent literature has focused on using SD, mainly through analyzing PSV and RI. In this regard, SD offers a significant advantage over CD and PD, namely the ability to obtain objective quantitative measurements, which could potentially reduce interobserver variability inherent to ultrasound examinations, particularly when using CD or PD modes. Regarding the discriminative capacity of DUS to distinguish NCAA from CAA, the available evidence is limited and currently markedly inferior to that reported for the diagnosis of AA versus a CG. This is a significant limitation, given that the potential presence of selection bias must be assumed in all cases.

An additional consideration is operator expertise. Most of the included studies were performed in academic centers, and examinations were conducted by experienced pediatric or abdominal radiologists, with some explicitly requiring more than five years of prior expertise in appendiceal US. Other reports indirectly underscored this dependence by restricting DUS examinations to working hours when trained radiologists were available. However, we did not identify any study that formally compared diagnostic performance stratified by operator expertise. This represents an important gap in the evidence and highlights a relevant avenue for future research.

Another relevant aspect is the lack of experience with the normal Doppler imaging appearance of the cecal appendix. This represents a significant limitation, as distinguishing between normal and pathological findings is critical for accurately characterizing the diagnostic performance of DUS in AA. It should also be considered that although the equipment used in the earlier studies was technologically more primitive and therefore less sensitive, it was reasonable to interpret a positive Doppler signal as pathological at that time. However, this concept likely requires re-evaluation given the greater sensitivity of current US machines.

Regarding study design, most studies were prospective with consecutive patient recruitment. However, a significant number of studies exhibited a potential risk of selection bias, as many included only patients in whom the appendix was visualized on grayscale US, and, in several cases, specifically those with borderline sonographic findings for AA diagnosis (e.g., non-compressible appendices or those measuring 6–8 mm). On the one hand, this represents an advantage, as the overall diagnostic performance of the tool is assessed in a population where diagnostic uncertainty is frequent, such as in cases of borderline visualized appendices. On the other hand, it must be noted that the diagnostic performance data provided in these studies may not reflect the general population of patients undergoing primary US for suspected AA, as cases without appendiceal visualization were systematically excluded in some studies. Considering the significant rate of non-visualization of the appendix reported in recent series, we believe that (1) the overall diagnostic performance of DUS in AA is likely overestimated in these studies, but on the other hand (2) this tool demonstrates potential diagnostic utility specifically in cases where the appendix is positively visualized, including those with borderline sonographic criteria.

Many retrospective studies relied on the retrospective review of static images or videotapes of examinations originally performed by other radiologists. We believe this represents a significant limitation and should be considered when evaluating the diagnostic performance reported in these studies. Additionally, the retrospective nature of these studies introduces essential limitations, such as (1) the lack of an accurate epidemiological representation of the prevalence of AA and its distribution by age and sex (for example, several studies report a disproportionately higher number of female patients, despite AA being a condition with a slight male predominance) [24]. Concerning the geographic distribution of the studies included, it is sufficiently broad not to limit the extrapolation of the results of this work.

Diagnostic odds ratios (DORs) across individual studies (Supplementary File 5) showed substantial variability, with some suggesting near-perfect performance but accompanied by wide confidence intervals. This asymmetry, with much wider upper bounds, is expected due to the log transformation applied during analysis and reflects the natural variability common to diagnostic accuracy studies [50]. Although DOR is less intuitive for clinical interpretation than sensitivity, specificity, or likelihood ratios, it remains a valuable composite measure of test performance. Overall, these findings highlight the influence of methodological and contextual factors not fully captured by subgroup analyses and underscore the need for standardized thresholds and prospective multicenter validation.

As a final remark, and as briefly mentioned in the Introduction, considering the rising prevalence of obesity in both pediatric and adult populations, it is essential to note that body habitus is a significant determinant of appendix visualization. Large retrospective and prospective series consistently show that overweight and obese patients are substantially more likely to have nondiagnostic examinations: visualization rates decrease from 85.7% in underweight children to only 29.3% in those with BMI z-scores >2 [51]; in another emergency cohort, 68% of nondiagnostic studies occurred in overweight patients, with BMI >85th percentile conferring nearly a five-fold increased odds of nondiagnostic results [52]; and a third prospective pediatric series confirmed reduced diagnostic accuracy in obese children (83% versus 93% in lean counterparts) [53]. Beyond obesity, anatomical factors such as retrocecal or pelvic locations further hinder sonographic identification. These findings underscore that patient habitus and appendix location remain critical constraints on ultrasound performance in suspected appendicitis.

The present study has key strengths, including its rigorous methodology aligned with PRISMA-DTA and the Cochrane Handbook for Diagnostic Test Accuracy Reviews, as well as the advanced meta-analytical models applied. *However, it has significant limitations: 1) the potential selection (spectrum) bias in most articles. Many of the included studies restricted their analyses to patients in whom the appendix could be visualized, or even to those with borderline grayscale findings. This approach artificially enriches the study population and likely inflates diagnostic accuracy estimates, since it systematically excludes the large proportion of patients with non-visualized appendices. Given the well-documented frequency of nondiagnostic examinations—particularly in obese patients or when the appendix is located in retrocecal or pelvic positions—our pooled estimates should be interpreted as applicable only to the subset of patients in whom the appendix is clearly identified, 2) the limitations inherent to the inferential statistical procedures used, 3) the small sample size and the retrospective nature of some of the included studies, 4) the high heterogeneity observed in some of the DTA meta-analytic models conducted, 5) the high heterogeneity in the CG definition, 6) the hierarchical model assessing "overall Doppler modalities" combines different Doppler techniques (CD, PD, CEPD, and SD) from overlapping study populations. This violates the assumption of independent observations and may lead to inflated estimates of precision. For this reason, this combined model should be interpreted strictly as exploratory. In contrast, separate models by modality provide more reliable estimates, 7) the pooled AUC estimates derived from hierarchical models extend beyond the empirical ROC space of the included studies, representing a model-based extrapolation rather than an observed summary measure. Consequently, AUC values should be interpreted cautiously, as they may overestimate diagnostic performance, especially in the presence of heterogeneity or limited study numbers, 8) Although meta-regressions were pre-specified a priori, their inherently exploratory nature warrants cautious interpretation, 9) in the study by Aydin et al. (2019), the reported 2×2 contingency table was internally inconsistent. After failing to obtain clarification from the original authors, we adopted the most plausible interpretation, inverting the reported cells. While this unverifiable decision introduces a potential source of bias, the uncorrected data would have yielded a diagnostic odds ratio close to zero—suggesting a useless test—completely at odds with the broader literature on color Doppler. By contrast, the corrected data align with previously reported diagnostic performance and are thus more biologically and clinically plausible. Nevertheless, this intervention must be acknowledged as a critical limitation, and the robustness of our conclusions should be interpreted with this caveat in mind, and 10) Deeks’ funnel plot asymmetry test was statistically significant for the Color Doppler model (p=0.08), indicating potential small-study effects or publication bias.

With the current evidence, DUS has not proven to be a reliable tool for differentiating between CAA and NCAA. Therefore, its use cannot be recommended for this specific purpose. Nevertheless, it remains an interesting avenue for future research. On the other hand, when the cecal appendix is adequately visualized, DUS demonstrates high diagnostic accuracy for confirming or ruling out acute appendicitis. Given its non-invasive nature and robust diagnostic performance, DUS holds promise as a vital diagnostic tool for the diagnosis of acute appendicitis. However, the limitations of this review, which are inherent to the primary studies, warrant further validation through large, well-designed multicenter studies.

## Supplementary Materials

**Supplementary File 1.** PRISMA-DTA checklist

**Supplementary File 2.** Full search strategy.

**Supplementary File 3.** Inclusion and exclusion criteria.

**Supplementary File 4.** <u>Above</u>: Exploratory, non-independent, DTA meta-analysis for overall Doppler (AA versus CG). HSROC curve. <u>Bottom</u>: Forest plot of the exploratory, non-independent, DTA meta-analysis performed for overall Doppler (AA versus CG).

**Supplementary File 5.** Dataset of included studies used for the review.

## Data Availability

STATEMENT OF AVAILABILITY OF THE DATA USED DURING THE SYSTEMATIC REVIEW:
All data used for the meta-analytical models are available in the accompanying supplementary dataset file.

