## Supplementary File 1 for "Diagnostic Performance of Doppler Ultrasound for Acute Appendicitis: A Systematic Review and Diagnostic Test Accuracy Meta-Analysis"

| **Section/topic** | **#** | **PRISMA-DTA Checklist Item** | **Reported on page #** |
| --- | --- | --- | --- |
| **TITLE / ABSTRACT** | | |  |
| Title | 1 | Identify the report as a systematic review (+/- meta-analysis) of diagnostic test accuracy (DTA) studies. | 1. Title. |
| Abstract | 2 | Abstract: See PRISMA-DTA for abstracts. | 3. Abstract. |
| **INTRODUCTION** | | |  |
| Rationale | 3 | Describe the rationale for the review in the context of what is already known. | 5,6. Introduction. |
| Clinical role of index test | D1 | State the scientific and clinical background, including the intended use and clinical role of the index test, and if applicable, the rationale for minimally acceptable test accuracy (or minimum difference in accuracy for comparative design). | 5,6. Introduction. |
| Objectives | 4 | Provide an explicit statement of question(s) being addressed in terms of participants, index test(s), and target condition(s). | 6,7. Introduction. |
| **METHODS** | | |  |
| Protocol and registration | 5 | Indicate if a review protocol exists, if and where it can be accessed (e.g., Web address), and, if available, provide registration information including registration number. | 3. Abstract.  7. Literature search and selection. |
| Eligibility criteria | 6 | Specify study characteristics (participants, setting, index test(s), reference standard(s), target condition(s), and study design) and report characteristics (e.g., years considered, language, publication status) used as criteria for eligibility, giving rationale. | 7. Literature search and selection. Supplementary File 3. |
| Information sources | 7 | Describe all information sources (e.g., databases with dates of coverage, contact with study authors to identify additional studies) in the search and date last searched. | 7. Literature search and selection. |
| Search | 8 | Present full search strategies for all electronic databases and other sources searched, including any limits used, such that they could be repeated. | 7. Literature search and selection. Supplementary File 2. |
| Study selection | 9 | State the process for selecting studies (i.e., screening, eligibility, included in systematic review, and, if applicable, included in the meta-analysis). | 7. Literature search and selection. Figure 1. |
| Data collection process | 10 | Describe method of data extraction from reports (e.g., piloted forms, independently, in duplicate) and any processes for obtaining and confirming data from investigators. | 7,8. Data extraction and synthesis. |
| Definitions for data extraction | 11 | Provide definitions used in data extraction and classifications of target condition(s), index test(s), reference standard(s) and other characteristics (e.g. study design, clinical setting). | 8,9. Data extraction and synthesis. |
| Risk of bias and applicability | 12 | Describe methods used for assessing risk of bias in individual studies and concerns regarding the applicability to the review question. | 7,8. Quality assessment. Figure 2. |
| Diagnostic accuracy measures | 13 | State the principal diagnostic accuracy measure(s) reported (e.g. sensitivity, specificity) and state the unit of assessment (e.g. per-patient, per-lesion). | 7,8. Data extraction and synthesis. 9,10. Meta-analysis |
| Synthesis of results | 14 | Describe methods of handling data, combining results of studies and describing variability between studies. This could include, but is not limited to: a) handling of multiple definitions of target condition. b) handling of multiple thresholds of test positivity, c) handling multiple index test readers, d) handling of indeterminate test results, e) grouping and comparing tests, f) handling of different reference standards | 7,8. Data extraction and synthesis. 9,10. Meta-analysis |

Page 1 of 2

| **Section/topic** | **#** | **PRISMA-DTA Checklist Item** | **Reported on page #** |
| --- | --- | --- | --- |
| Meta-analysis | D2 | Report the statistical methods used for meta-analyses, if performed. | 9-11. Meta-analysis, diagnostic test accuracy meta-analysis, publication bias and small-study effects assessment. |
| Additional analyses | 16 | Describe methods of additional analyses (e.g., sensitivity or subgroup analyses, meta-regression), if done, indicating which were pre-specified. | 9-11. Meta-analysis, diagnostic test accuracy meta-analysis, publication bias and small-study effects assessment. |
| **RESULTS** | | |  |
| Study selection | 17 | Provide numbers of studies screened, assessed for eligibility, included in the review (and included in meta-analysis, if applicable) with reasons for exclusions at each stage, ideally with a flow diagram. | 11,12. Results |
| Study characteristics | 18 | For each included study provide citations and present key characteristics including: a) participant characteristics (presentation, prior testing), b) clinical setting, c) study design, d) target condition definition, e) index test, f) reference standard, g) sample size, h) funding sources | 11-14. Results. Sociodemographic characteristics. |
| Risk of bias and applicability | 19 | Present evaluation of risk of bias and concerns regarding applicability for each study. | 11,12. Results |
| Results of individual studies | 20 | For each analysis in each study (e.g. unique combination of index test, reference standard, and positivity threshold) report 2x2 data (TP, FP, FN, TN) with estimates of diagnostic accuracy and confidence intervals, ideally with a forest or receiver operator characteristic (ROC) plot. | 11-20. Results. Table 1. |
| Synthesis of results | 21 | Describe test accuracy, including variability; if meta-analysis was done, include results and confidence intervals. | 3. Abstract. 11-20. Results. Figures 3-6. |
| Additional analysis | 23 | Give results of additional analyses, if done (e.g., sensitivity or subgroup analyses, meta-regression; analysis of index test: failure rates, proportion of inconclusive results, adverse events). | 14-15. Meta-analysis |
| **DISCUSSION** | | |  |
| Summary of evidence | 24 | Summarize the main findings including the strength of evidence. | 20-23. Discussion |
| Limitations | 25 | Discuss limitations from included studies (e.g. risk of bias and concerns regarding applicability) and from the review process (e.g. incomplete retrieval of identified research). | 23. Discussion |
| Conclusions | 26 | Provide a general interpretation of the results in the context of other evidence. Discuss implications for future research and clinical practice (e.g. the intended use and clinical role of the index test). | 23. Discussion |
| **FUNDING** | | |  |
| Funding | 27 | For the systematic review, describe the sources of funding and other support and the role of the funders. | 1,2. Title page. |

*Adapted From:* McInnes MDF, Moher D, Thombs BD, McGrath TA, Bossuyt PM, The PRISMA-DTA Group (2018). Preferred Reporting Items for a Systematic Review and Meta-analysis of Diagnostic Test Accuracy Studies: The PRISMA-DTA Statement. JAMA. 2018 Jan 23;319(4):388-396. doi: 10.1001/jama.2017.19163.

Page 2 of 2
