## Supplementary File 2 for "Diagnostic Performance of Doppler Ultrasound for Acute Appendicitis: A Systematic Review and Diagnostic Test Accuracy Meta-Analysis"

**Supplementary File 2: Search Strategy**

We will use a two-part search strategy to identify studies that meet the inclusion criteria and were published between January 1, 1990, and April 22, 2025:

- We will search bibliographic databases in the fields of medicine and public health using a comprehensive search strategy targeting:
  - The diagnostic performance of Doppler ultrasound in adult and pediatric acute appendicitis;
  - The ability of Doppler ultrasound to discriminate between complicated and uncomplicated acute appendicitis in both populations.
- We will also conduct backward snowballing by screening the reference lists of all primary studies included in the review and those of relevant previously published systematic reviews.

The following electronic databases will be searched: PubMed, Web of Science, Scopus, and Ovid MEDLINE.

**Search strategy**

**PUBMED =** 91

((appendicitis[MeSH Terms] OR appendicitis[Title/Abstract] OR "acute appendicitis"[Title/Abstract]) AND (("Doppler"[Title/Abstract] OR "color flow"[Title/Abstract] OR "blood flow"[Title/Abstract] OR "hyperemia"[Title/Abstract] OR "vascular signal"[Title/Abstract] OR "flow signal"[Title/Abstract] OR "increased flow"[Title/Abstract] OR "color mapping"[Title/Abstract] OR "color sonography"[Title/Abstract] OR "flow detection"[Title/Abstract])) AND (diagnosis[Title/Abstract] OR diagnostic[Title/Abstract]) NOT (review[Publication Type] OR "systematic review"[Title])

**WoS =** 192

TS=(appendicitis OR "acute appendicitis")

AND TS=("Doppler ultrasound" OR "color Doppler" OR "power Doppler" OR "spectral Doppler" OR "Doppler sonography" OR "Doppler ultrasonography" OR "Doppler US"

OR "color flow" OR "blood flow" OR "vascular flow" OR "hyperemia" OR "increased flow" OR "color mapping" OR "flow detection" OR "vascular signal") AND TS=(diagnosis OR diagnostic OR "diagnostic imaging") NOT TS=("systematic review" OR "review")

**Scopus =** 92

TITLE-ABS-KEY ( appendicitis AND ( "Doppler ultrasound" OR "color Doppler" OR "spectral Doppler" OR "contrast-enhanced Doppler" OR "resistance index" ) AND ( diagnosis OR specificity OR sensitivity OR "diagnostic performance" OR "equivocal Alvarado score" OR "borderline appendix" ) ) AND ( EXCLUDE ( DOCTYPE , "re" ) OR EXCLUDE ( DOCTYPE , "le" ) )

**OVID MEDLINE =** 30

(exp Appendicitis/ OR appendicitis.tw. OR "acute appendicitis".tw.) AND

(exp Ultrasonography, Doppler/ OR "doppler ultrasound".tw. OR "color doppler".tw. OR "spectral doppler".tw. OR "resistance index".tw.) AND

("diagnostic performance".tw. OR specificity.tw. OR sensitivity.tw. OR "equivocal Alvarado score".tw. OR "borderline appendix".tw.)
