## Supplementary File 3 for "Diagnostic Performance of Doppler Ultrasound for Acute Appendicitis: A Systematic Review and Diagnostic Test Accuracy Meta-Analysis"

**Supplementary File 3: Inclusion and Exclusion Criteria**

**Inclusion Criteria**

- Prospective or retrospective original observational clinical studies evaluating the diagnostic performance of any modality of ultrasonographic Doppler flow (e.g., color, power, spectral), performed by any examiner, for diagnosing acute appendicitis and/or discriminating between complicated and uncomplicated appendicitis in both adult and pediatric populations, when compared to the reference standard (histopathological analysis of the resected cecal appendix).

**Exclusion Criteria**

- Case reports.
- Duplicate or overlapping studies.
- Retracted studies.
- Reviews, systematic reviews, or clinical guidelines.
- Letters to the editor and editorials.
- Articles published in languages other than English or Spanish.
- Studies with no surgical intervention.
- Studies that do not include the population of interest.
- Studies conducted exclusively in immunocompromised patients.
- Studies involving patients with metastatic or invasive abdominal neoplastic disease.
- Studies involving patients with hematological disorders.
