## Supplementary figures and images for "Diagnostic Performance of Doppler Ultrasound for Acute Appendicitis: A Systematic Review and Diagnostic Test Accuracy Meta-Analysis"

### Supplementary File 4

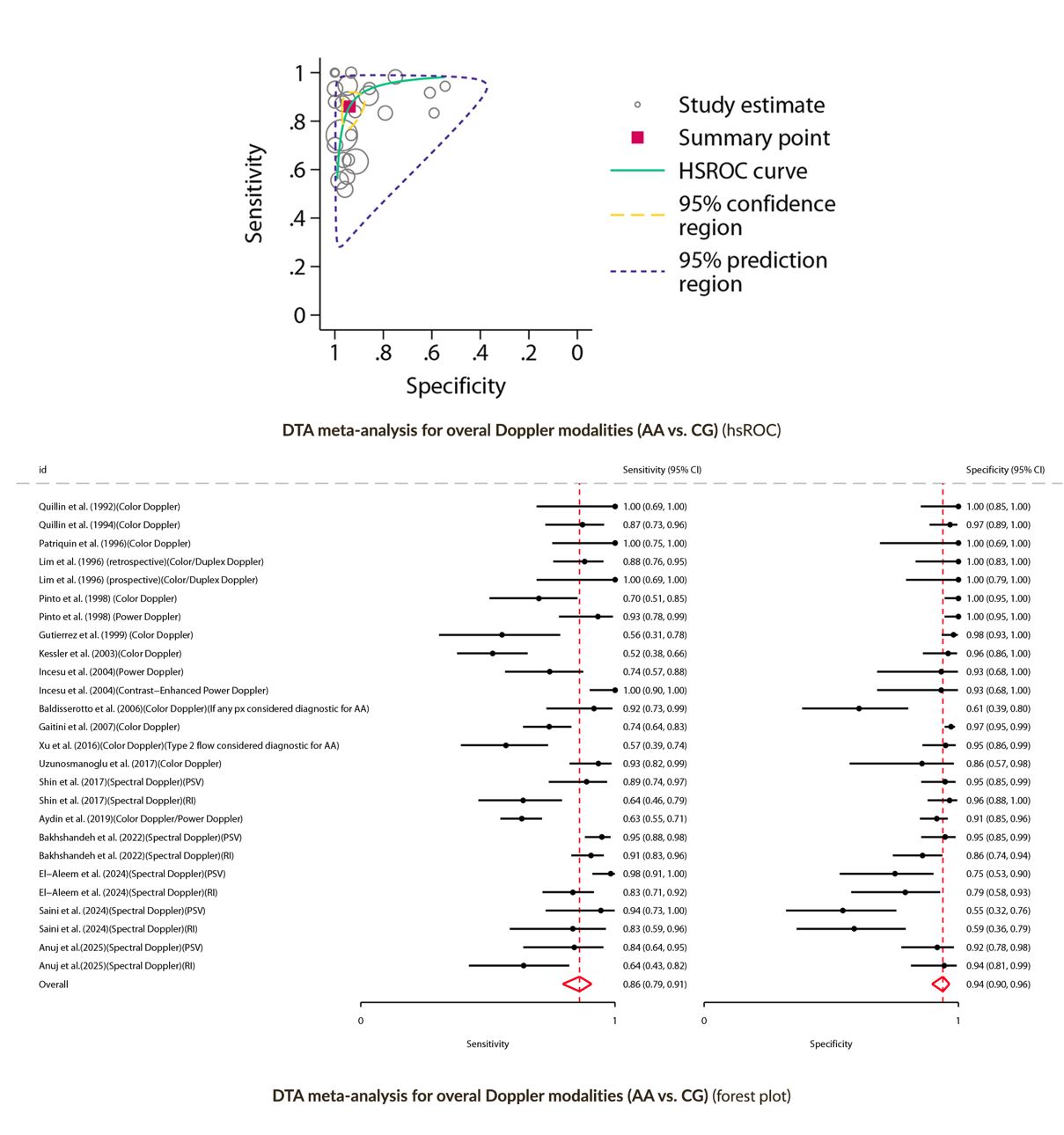
